# Hydroxychloroquine for prevention of COVID-19 mortality: a population-based cohort study

**DOI:** 10.1101/2020.09.04.20187781

**Authors:** Christopher T Rentsch, Nicholas J DeVito, Brian MacKenna, Caroline E Morton, Krishnan Bhaskaran, Jeremy P Brown, Anna Schultze, William J Hulme, Richard Croker, Alex J Walker, Elizabeth J Williamson, Chris Bates, Seb Bacon, Amir Mehrkar, Helen J Curtis, David Evans, Kevin Wing, Peter Inglesby, Rohini Mathur, Henry Drysdale, Angel YS Wong, Helen I McDonald, Jonathan Cockburn, Harriet Forbes, John Parry, Frank Hester, Sam Harper, Liam Smeeth, Ian J Douglas, William G Dixon, Stephen JW Evans, Laurie Tomlinson, Ben Goldacre

## Abstract

**Background:** Hydroxychloroquine has been shown to inhibit severe acute respiratory syndrome coronavirus 2 (SARS-CoV-2) in vitro, but early clinical studies found no benefit treating patients with coronavirus disease 2019 (COVID-19). We set out to evaluate the effectiveness of hydroxychloroquine for prevention, as opposed to treatment, of COVID-19 mortality.

**Methods:** We pre-specified and conducted an observational, population-based cohort study using national primary care data and linked death registrations in the OpenSAFELY platform, representing 40% of the general population in England. We used Cox regression to estimate the association between ongoing routine hydroxychloroquine use prior to the COVID-19 outbreak in England and risk of COVID-19 mortality among people with rheumatoid arthritis (RA) or systemic lupus erythematosus (SLE). Model adjustment was informed by a directed acyclic graph.

**Results:** Of 194,637 patients with RA or SLE, 30,569 (15.7%) received ≥ 2 prescriptions of hydroxychloroquine in the six months prior to 1 March 2020. Between 1 March 2020 and 13 July 2020, there were 547 COVID-19 deaths, 70 among hydroxychloroquine users. Estimated standardised cumulative COVID-19 mortality was 0.23% (95% CI 0.18–0.29) among users and 0.22% (95% CI 0.20–0.25) among non-users; an absolute difference of 0.008% (95% CI –0.051-0.066). After accounting for age, sex, ethnicity, use of other immunuosuppressives, and geographic region, no association with COVID-19 mortality was observed (HR 1.03, 95% CI 0.80–1.33). We found no evidence of interactions with age or other immunosuppressives. Quantitative bias analyses indicated observed associations were robust to missing information regarding additional biologic treatments for rheumatological disease. We observed similar associations with the negative control outcome of non-COVID-19 mortality.

**Conclusion:** We found no evidence of a difference in COVID-19 mortality among patients who received hydroxychloroquine for treatment of rheumatological disease prior to the COVID-19 outbreak in England.

**Research in context:** *Evidence before this study:* Published trials and observational studies to date have shown no evidence of benefit of hydroxychloroquine as a treatment for hospitalised patients who already have COVID-19. A separate question remains: whether routine ongoing use of hydroxychloroquine in people without COVID-19 protects against new infections or severe outcomes. We searched MEDLINE/PubMed for pharmacoepidemiological studies evaluating hydroxychloroquine for prevention of severe COVID-19 outcomes. The keywords “hydroxychloroquine AND (COVID OR coronavirus OR SARS-CoV-2) AND (prophyl* OR prevent*) AND (rate OR hazard OR odds OR risk)” were used and results were filtered to articles from the last year with abstracts available. 109 papers were identified for screening; none investigated pre-exposure prophylactic use of hydroxychloroquine for prevention of severe COVID-19 outcomes. Clinical trials of prophylactic use of hydroxychloroquine are ongoing; however, the largest trial does not expect to meet recruitment targets due to “…unjustified extrapolation and exaggerated safety concerns together with intense politicisation and negative publicity.” In the absence of reported clinical trials, evidence can be generated from real-world data to support the need for randomised clinical trials.

*Added value of this study:* In this cohort study representing 40% of the population of England, we investigated whether routine use of hydroxychloroquine prior to the COVID-19 outbreak prevented COVID-19 mortality. Using robust pharmacoepidemiological methods, we found no evidence to support a substantial benefit of hydroxychloroquine in preventing COVID-19 mortality. At the same time, we have shown no significant harm, and this generates the equipoise to justify continuing randomised trials. We have demonstrated in this study that it is feasible to address specific hypotheses about medicines in a rapid and transparent manner to inform interim clinical decision making and support the need for large-scale, randomised trial data.

*Implications of all the available evidence:* This is the first study to investigate the ongoing routine use of hydroxychloroquine and risk of COVID-19 mortality in a general population. While we found no evidence of any protective benefit, due to the observational nature of the study, residual confounding remains a possibility. Completion of trials for prevention of severe outcomes is warranted, but prior to the completion of these, we found no evidence to support the use of hydroxychloroquine for prevention of COVID-19 mortality.

## Background

Hydroxychloroquine, a commonly used conventional synthetic disease-modifying antirheumatic drug (sDMARD), is indicated for treatment of rheumatoid arthritis (RA) and systemic lupus erythematosus (SLE).^1^ Early in the severe acute respiratory syndrome coronavirus 2 (SARS-CoV-2) pandemic, it was suggested that hydroxychloroquine may benefit the treatment and prevention of coronavirus disease 2019 (COVID-19).^2–4^ Hydroxychloroquine has since been investigated in numerous clinical trials^5–9^ and observational cohorts^10–12^ with no evidence of therapeutic efficacy in treatment of hospitalised patients with symptomatic COVID-19.

Evaluations of the effectiveness of hydroxychloroquine for prevention, as opposed to treatment, of SARS-CoV-2 infection or severe COVID-19 outcomes are limited.^13^ One randomised, controlled trial examining hydroxychloroquine as post-exposure prophylaxis did not demonstrate significant benefit in preventing SARS-CoV-2 infection, though uncertainty in results could not exclude possible benefit.^14^ Other trials for the prevention of COVID-19 outcomes are ongoing.^5^ In this large population-based cohort study, we examined whether ongoing routine hydroxychloroquine use prior to the outbreak in England was associated with lower risk of COVID-19 mortality.

## Methods

### Study design and population

We conducted an observational cohort study using electronic health record (EHR) data from primary care practices using The Phoenix Partnership (TPP) software linked to Office for National Statistics (ONS) death registrations through OpenSAFELY. This is a data analytics platform developed during the COVID-19 pandemic to allow near real-time analysis of pseudonymised primary care patient records at scale, covering approximately 40% of the population in England, operating within the EHR vendor’s highly secure data centre.^15,16^ Pseudonymised structured data include demographics, medications prescribed from primary care, diagnoses, and laboratory measures. Details on Information Governance of the OpenSAFELY platform can be found in the **Supplementary Appendix**.

We included all adults aged ≥ 18 years registered with a general practice for ≥1 year on 1 March 2020 (index date) with information on age, sex, and deprivation. Within this source population, we identified people who were diagnosed with RA or SLE ≥6 months prior to the index date and therefore had indication for hydroxychloroquine use prior to the outbreak in England.^17^ We studied people with these conditions to minimise the potential for confounding by indication when estimating the effectiveness of hydroxychloroquine use rather than investigate how to prevent severe COVID-19 in this population.

### Exposure, outcome and follow-up

The exposure of interest was regular use of hydroxychloroquine (≥2 prescriptions in the 6 months prior to index, “users”) compared to no regular use of hydroxychloroquine (“non-users”). The primary outcome was COVID-19 mortality, defined by the presence of ICD-10 codes U07.1 (“COVID-19, virus identified”) or U07.2 (“COVID-19, virus not identified”) on the death certificate.^18^ We followed people from index date until earliest of: date of death, or seven days prior to last date of availability of ONS mortality data to account for reporting lag. People not exposed to hydroxychloroquine before index date were censored if prescribed it during follow-up. Study design is depicted in **eFigure 1 in Supplementary Appendix**.

**Figure 1:**
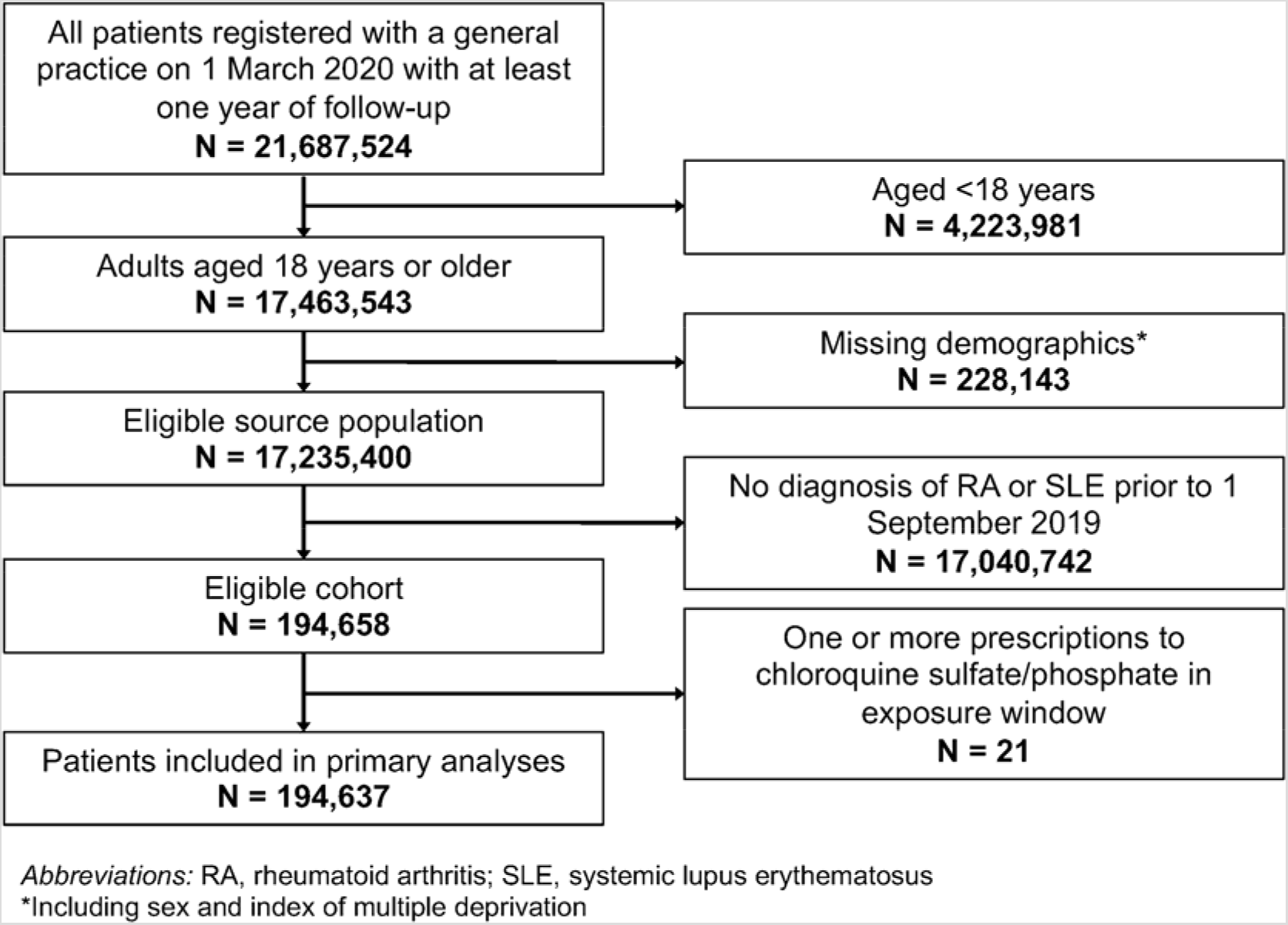
Flow chart

### Covariates

Potential determinants of regular hydroxychloroquine use and COVID-19 mortality were identified by reviewing existing literature and through discussions with clinicians. As this was a study of prevalent hydroxychloroquine users, we included determinants that may have influenced the initial choice of treatment and whether people remained on treatment. The full list of pre-specified variables included age, sex, ethnicity, index of multiple deprivation quintile, other immunosuppressives (other sDMARDs, oral corticosteroids), smoking status, prescribed non-steroidal anti-inflammatory drugs (NSAIDs), body mass index, hypertension, diabetes severity as measured by diagnostic codes and glycated haemoglobin (HbA1c), heart disease, liver disease, respiratory disease excluding asthma, kidney disease as measured by estimated glomerular filtration rate (eGFR), stroke, dementia, cancer, and influenza vaccination in 2019/20 season. Our methodology for creating codelists has been previously described.^15^ We developed a directed acyclic graph (DAG) to identify the minimal set of covariates to adjust for the hypothesised confounding structure, which included age, sex, ethnicity, geographic region, and other immunosuppressives (see **eFigure 2 in Supplementary Appendix**).

**Figure 2:**
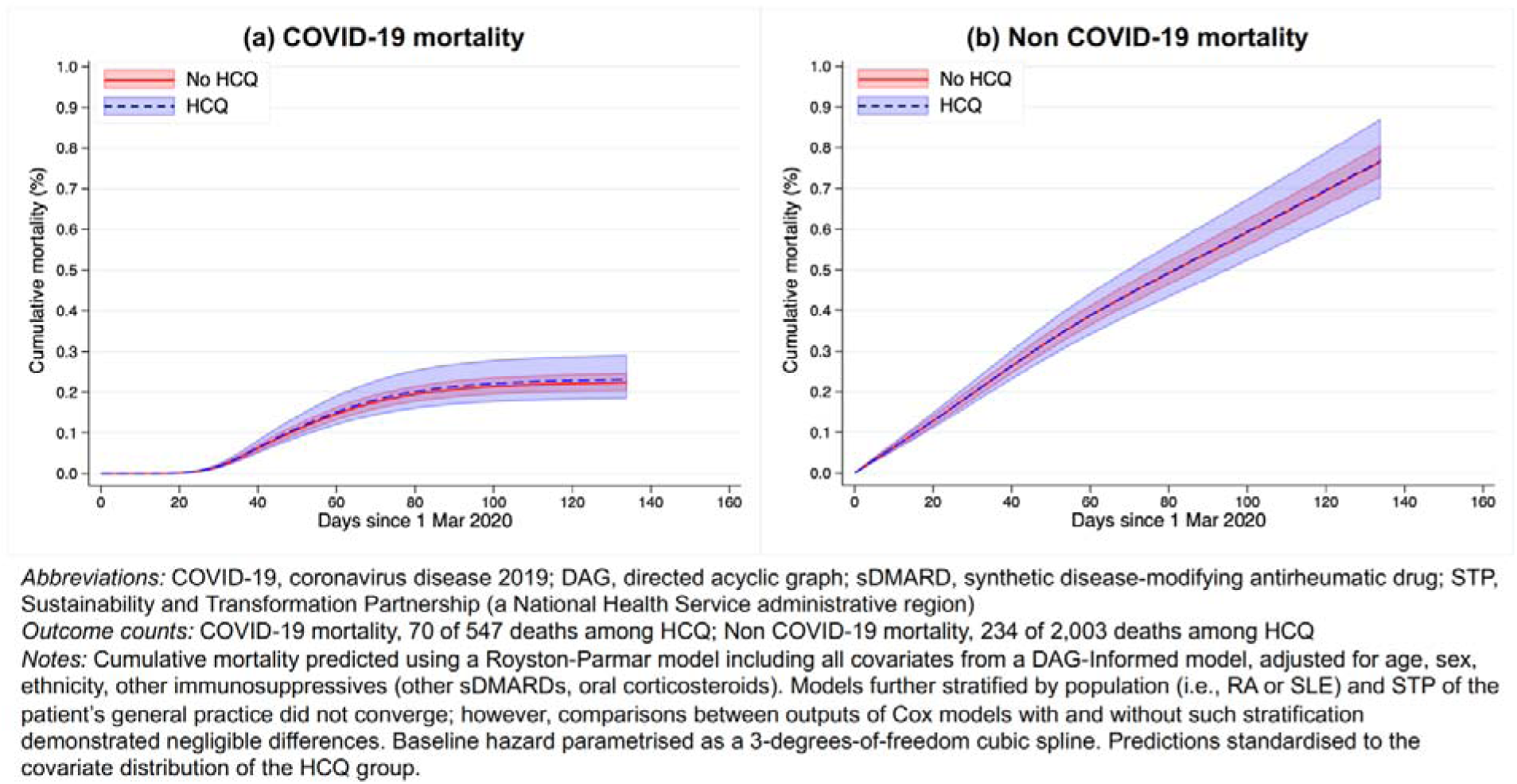
Cumulative mortality by hydroxychloroquien (HCQ) use among 194,637 patients with rheumatoid arthritics (RA) or systemic lupus erythematosus (SLE)

### Statistical methods

Patient characteristics were summarised using descriptive statistics, stratified by exposure status. We used Cox regression models with days since the index date as the timescale to estimate hazard ratios (HRs) and 95% confidence intervals (CIs) for the association between regular hydroxychloroquine use and COVID-19 mortality. The competing risk of death from causes other than COVID-19 was accounted for by censoring non COVID-19 deaths; our analysis therefore estimated cause-specific hazards.^19^ We sequentially adjusted for sex and age using restricted cubic splines; for the minimal adjustment set informed by the DAG; and finally extended for all extracted covariates listed above. Models were stratified by an indicator variable denoting patient population(i.e., RA or SLE) and geographic region. Multiple imputation (10 imputations) was used to account for missing ethnicity for 23% of the sample, with the imputation model including all extracted covariates and an indicator for the outcome. Those with missing body mass index were assumed to be non-obese, and those with missing smoking data were assumed to be never-smokers; we did not use multiple imputation for these variables as they are expected to be missing not at random in UK primary care.^20^ Proportional hazards were checked by examining the Schoenfeld residuals over time.

We generated cumulative mortality curves standardised to adjust for different covariate distributions in the exposed group. First, a flexible parametric Royston-Parmar model with the same covariates as the DAG-Informed Cox model was fitted, with the baseline hazard modelled using a three degrees-of-freedom spline. The survival function was predicted from this model for every individual with regular hydroxychloroquine use and averaged to produce the curve for the exposed group. To produce the standardised comparison curve, the survival functions were predicted and averaged again for the same individuals, but with exposure set to zero. Patient population was included in the flexible parametric model as a binary indicator variable since the model could not converge with both patient population and geographic region as stratification variables. Comparisons between Cox and Royston-Parmar models can be found in the **Supplementary Appendix**.

We evaluated pre-specified interactions to determine whether the association between regular hydroxychloroquine use and COVID-19 mortality varied by age, exposure to other sDMARDs, oral corticosteroids, and NSAIDs. Two-sided p-values were calculated from Wald tests on interaction terms.

### Sensitivity analyses

We adjusted for ethnicity in a model excluding people with missing ethnicity and compared results with those from multiple imputation. In primary analyses, < 2% of the comparison group had one prescription of hydroxychloroquine in the six-month exposure window. We re-defined exposure as ≥ 1 prescription in the three months prior to the index date. We compared results from primary models stratified for patient population (RA or SLE) to a model that included patient population as a binary indicator variable as well as modelling each population separately.

We calculated bias-adjusted hazard ratios to evaluate how adjustment for biologic DMARDs (bDMARDs), including targeted synthetic DMARDs (tsDMARDs), which were not available for this analysis, may have produced different results under differing assumptions of prevalence and effect on COVID-19 mortality.^21^ Prevalence of bDMARDs in each exposure group was estimated from preexisting literature (18% among users and 21% among non-users);^22,23^ however, we also examined more extreme values of prevalence (see **Supplementary Appendix**). We assumed a range of potential associations between bDMARDs and COVID-19 mortality from 0.8 to 1.2.

We conducted analyses using non COVID-19 mortality as a negative control outcome, censoring at COVID-19 death. We hypothesized that If associations observed in primary analyses were due to confounding by indication, we would observe a similar association with non COVID-19 mortality.

### Software and reproducibility

Data management was performed using Python 3.8 and SQL, and analysis using Stata 16.1. All code for data management and analyses is at: https://github.com/opensafely/hydroxychloroquineresearch. All iterations of the pre-specified protocol are archived with version control at: https://github.com/opensafely/hydroxychloroquine-research/tree/master/protocol.

## Results

### Patient characteristics

We identified 194,637 people with RA or SLE at least six months prior to 1 March 2020 (i.e., index date) for analysis (**Figure 1**).

Of these, 30,569 (15.7%) demonstrated regular use of hydroxychloroquine in the six months prior to index date. Hydroxychloroquine users were younger (median: 63 years for users, 66 years for nonusers) and more likely to be female (76% women for users; 70% for non-users); other demographic characteristics between exposure groups were broadly similar (**Table 1**). Hydroxychloroquine users were more likely to be on other sDMARDs (52% vs 34%), oral corticosteroids (23% vs 16%), and NSAIDs (22% vs 16%). Distributions of characteristics in RA and SLE populations are shown in **eTable 1** and **eTable 2**, respectively.

**Table 1.** Demographic and clinical characteristics of 194,637 patients with rheumatoid arthritis or systemic lupus erythematosus (SLE)

|  | Total | No HCQ | HCQ |
| --- | --- | --- | --- |
| Sample size, n | 194,637 (100.0) | 164,068 (100.0) | 30,569 (100.0) |
| **Population** |  |  |  |
| Rheumatoid arthritis | 167,874 (86.2) | 144,151 (87.9) | 23,723 (77.6) |
| SLE | 26,763 (13.8) | 19,917 (12.1) | 6,846 (22.4) |
| **Demographics** |  |  |  |
| Age, years |  |  |  |
| Median (IQR) | 66 (54-75) | 66 (55-76) | 63 (53-72) |
| 18-39 | 13,709 (7.0) | 11,433 (7.0) | 2,276 (7.4) |
| 40-49 | 19,438 (10.0) | 15,829 (9.6) | 3,609 (11.8) |
| 50-59 | 37,086 (19.1) | 30,457 (18.6) | 6,629 (21.7) |
| 60-69 | 45,699 (23.5) | 37,726 (23.0) | 7,973 (26.1) |
| 70-79 | 49,238 (25.3) | 42,090 (25.7) | 7,148 (23.4) |
| ≥80 | 29,467 (15.1) | 26,533 (16.2) | 2,934 (9.6) |
| Sex |  |  |  |
| Female | 138,440 (71.1) | 115,106 (70.2) | 23,334 (76.3) |
| Male | 56,197 (28.9) | 48,962 (29.8) | 7,235 (23.7) |
| Ethnicity |  |  |  |
| White | 132,697 (68.2) | 112,367 (68.5) | 20,330 (66.5) |
| South Asian | 10,498 (5.4) | 8,502 (5.2) | 1,996 (6.5) |
| Black | 2,997 (1.5) | 2,425 (1.5) | 572 (1.9) |
| Mixed | 1,279 (0.7) | 1,005 (0.6) | 274 (0.9) |
| Other | 1,838 (0.9) | 1,508 (0.9) | 330 (1.1) |
| Missing | 45,328 (23.3) | 38,261 (23.3) | 7,067 (23.1) |
| Index of multiple deprivation |  |  |  |
| 1 (least deprived) | 38,968 (20.0) | 32,954 (20.1) | 6,014 (19.7) |
| 2 | 39,437 (20.3) | 33,351 (20.3) | 6,086 (19.9) |
| 3 | 38,942 (20.0) | 32,800 (20.0) | 6,142 (20.1) |
| 4 | 38,477 (19.8) | 32,402 (19.7) | 6,075 (19.9) |
| 5 (most deprived) | 38,813 (19.9) | 32,561 (19.8) | 6,252 (20.5) |
| Residence type |  |  |  |
| Rural | 45,656 (23.5) | 38,305 (23.3) | 7,351 (24.0) |
| Urban | 148,981 (76.5) | 125,763 (76.7) | 23,218 (76.0) |
| Body mass index, kg/m<sup>2</sup> |  |  |  |
| <18.5 | 4,372 (2.2) | 3,692 (2.3) | 680 (2.2) |
| 18.5-24.9 | 56,981 (29.3) | 48,051 (29.3) | 8,930 (29.2) |
| 25-29.9 | 61,870 (31.8) | 52,667 (32.1) | 9,203 (30.1) |
| 30-34.9 | 35,315 (18.1) | 29,652 (18.1) | 5,663 (18.5) |
| 35-39.9 | 14,999 (7.7) | 12,372 (7.5) | 2,627 (8.6) |
| ≥40 | 7,727 (4.0) | 6,156 (3.8) | 1,571 (5.1) |
| Missing | 13,373 (6.9) | 11,478 (7.0) | 1,895 (6.2) |
| Smoking status |  |  |  |
| Never | 74,184 (38.1) | 62,705 (38.2) | 11,479 (37.6) |
| Former | 92,432 (47.5) | 77,740 (47.4) | 14,692 (48.1) |
| Current | 27,411 (14.1) | 23,079 (14.1) | 4,332 (14.2) |
| Missing | 610 (0.3) | 544 (0.3) | 66 (0.2) |
| **Clinical conditions** |  |  |  |
| Diabetes |  |  |  |
| No diabetes | 159,830 (82.1) | 133,954 (81.6) | 25,876 (84.6) |
| Diabetes, HbA1c <7.5% | 22,713 (11.7) | 19,560 (11.9) | 3,153 (10.3) |
| Diabetes, HbA1c ≥7.5% | 8,998 (4.6) | 7,930 (4.8) | 1,068 (3.5) |
| Diabetes, missing HbA1c | 3,096 (1.6) | 2,624 (1.6) | 472 (1.5) |
| eGFR, ml/min/1.73m<sup>2</sup> |  |  |  |
| ≥60 | 133,371 (68.5) | 109,606 (66.8) | 23,765 (77.7) |
| 30-59 | 24,528 (12.6) | 21,153 (12.9) | 3,375 (11.0) |
| <30 | 1,944 (1.0) | 1,698 (1.0) | 246 (0.8) |
| Missing | 34,794 (17.9) | 31,611 (19.3) | 3,183 (10.4) |
| Heart disease | 30,609 (15.7) | 26,292 (16.0) | 4,317 (14.1) |
| Liver disease | 2,718 (1.4) | 2,227 (1.4) | 491 (1.6) |
| Respiratory disease (excluding asthma) | 26,680 (13.7) | 22,159 (13.5) | 4,521 (14.8) |
| Neurological condition | 12,718 (6.5) | 11,003 (6.7) | 1,715 (5.6) |
| Hypertension | 83,404 (42.9) | 71,117 (43.3) | 12,287 (40.2) |
| Cancer | 20,028 (10.3) | 17,144 (10.4) | 2,884 (9.4) |
| Immunosuppression | 2,969 (1.5) | 2,399 (1.5) | 570 (1.9) |
| Influenza vaccination 2019/20 | 122,295 (62.8) | 101,112 (61.6) | 21,183 (69.3) |
| **Other medications** |  |  |  |
| Other sDMARD | 71,523 (36.7) | 55,780 (34.0) | 15,743 (51.5) |
| Azithromycin | 948 (0.5) | 751 (0.5) | 197 (0.6) |
| Oral corticosteroid | 33,677 (17.3) | 26,792 (16.3) | 6,885 (22.5) |
| NSAID | 33,356 (17.1) | 26,686 (16.3) | 6,670 (21.8) |

*Abbreviations:* IQR, interquartile range; HbA1c, glycated haemoglobin; eGFR, estimated glomerular filtration rate; sDMARD, synthetic disease-modifying antirheumatic drug; NSAID, non-steroidal anti-inflammatory drug

### Univariable and multivariable regression

Between 1 March 2020 and 13 July 2020, there were 547 COVID-19 deaths among people with RA or SLE, 70 of which were among regular users of hydroxychloroquine. Estimated standardised cumulative mortality was 0.23% (95% CI 0.18–0.29) and 0.22% (95% CI 0.20–0.25) among users and non-users, respectively, at the end of follow-up (**Figure 2**). The absolute cumulative risk difference was 0.008% (95% CI –0.051, 0.066). In unadjusted analyses, regular users of hydroxychloroquine had a decreased risk of COVID-19 mortality (HR 0.78, 95% CI 0.60–1.00, **Figure 3**). After adjusting for age and sex, there was no longer any evidence of association (HR 1.08, 95% CI 0.84- 0.80–1.40). Additionally adjusting for variables identified in the DAG (HR 1.03, 95% CI 0.80– 1.33) or extending to all covariates (HR 1.03, 95% CI 0.80–1.33) did not alter conclusions. There was no evidence of interaction by age, exposure to other sDMARDs, oral corticosteroids, or NSAIDs (**eTable 3**).

**Figure 3:**
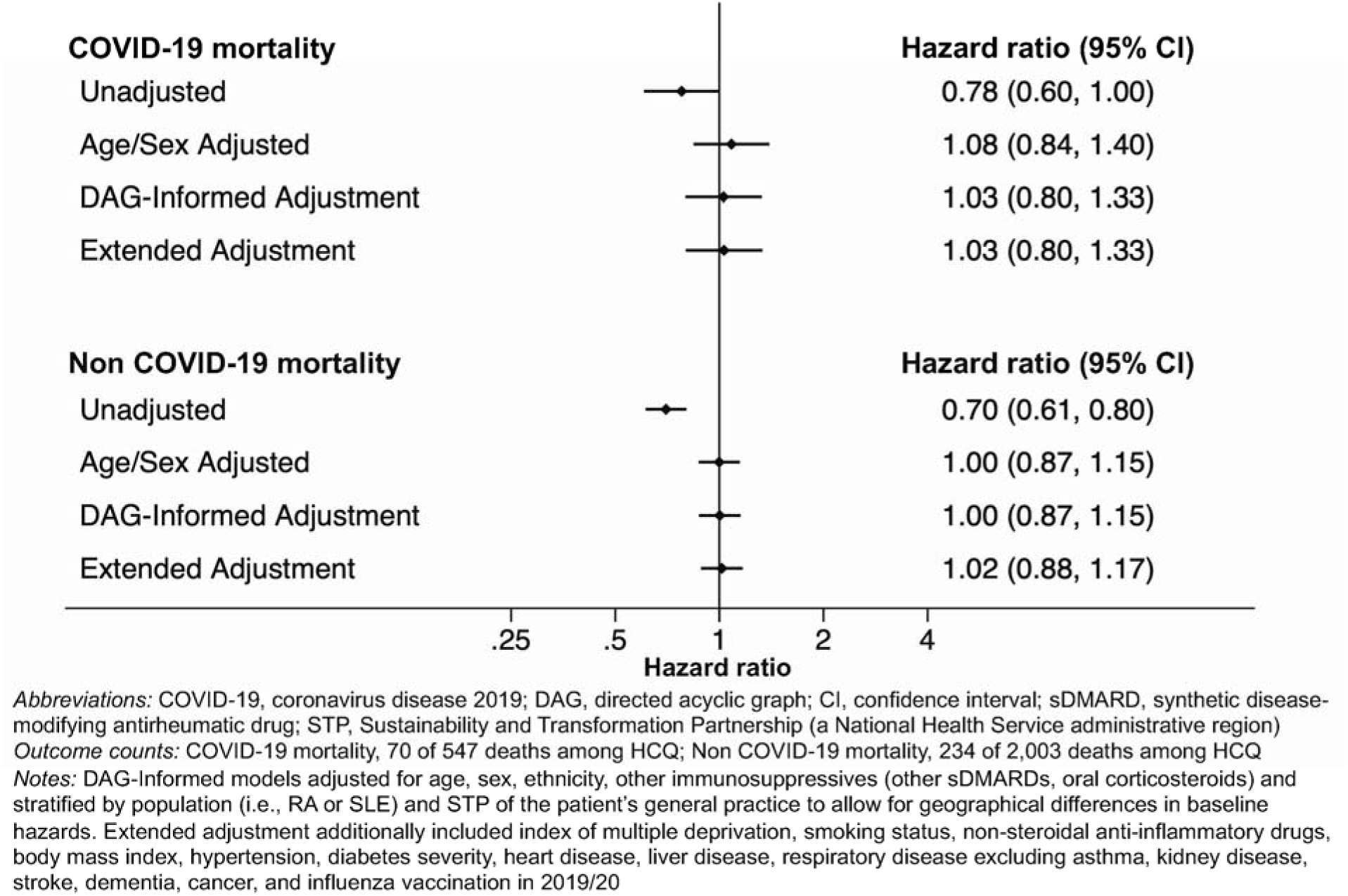
Comparison between hydroxychloroquien(HCQ)use and no HCQ use among 194,637 patients with rheumatoid arthritis(RA) or systemic lupus erythematosus (SLE)

### Sensitivity analyses

Results from all sensitivity analyses provided similar findings to primary analyses (**eTable 4**). In quantitative bias analyses, values of the bias-adjusted association ranged from HR 0.97 (95% CI 0.75–1.26) to HR 1.09 (95% CI 0.84–1.40) (**eTable 5**). Hydroxychloroquine use was not associated with the negative control outcome of non COVID-19 mortality after adjustment for age and sex (HR 1.00, 95% CI 0.87–1.15, **Figure 3**).

## Discussion

### Key findings

In this national, population-based study of hydroxychloroquine users, we found no evidence that hydroxychloroquine was associated with either a beneficial or harmful impact on COVID-19 mortality. The confidence intervals around the relative risk suggest that we could exclude substantial benefit, though a modest benefit or harm on a relative scale could not be ruled out. However, even if hydroxychloroquine provided a benefit, our results demonstrate a maximum absolute risk reduction of 0.05% in the context of an absolute risk of 0.22% of COVID-19 mortality among non-users. Taken together, our findings do not provide any strong support for a protective effect from ongoing routine hydroxychloroquine use as has been previously hypothesised. Our estimates were robust to multiple sensitivity analyses. We have demonstrated in this study that it is feasible to address specific hypotheses about medicines in a transparent manner in response to speculation, using OpenSAFELY, and to inform regulatory bodies decision making in the absence of high quality, randomised trial data. However, due to the observational nature of the study, a degree of uncertainty persists that can only be addressed through large-scale randomised trials.

### Comparison with other evidence

A randomised trial examining hydroxychloroquine for post-exposure prophylaxis did not demonstrate a significant benefit in preventing infection, although the findings were compatible with an absolute risk reduction as much as 7% in the context of an absolute risk of about 14% in the placebo arm.^14^ As we await the reporting of ongoing clinical trials of prophylactic use of hydroxychloroquine, related evidence of drug effectiveness among existing users can be generated from observational data. Previous investigations of hydroxychloroquine using observational data^24^ had limitations in their design and analysis,^25^ including relatively small sample sizes and focusing only on hospitalised patients, which may produce spurious associations.^26^

Numerous randomised trials^5–9^ have failed to find any clinical benefit of hydroxychloroquine for treatment of COVID-19. Studies to date have also not demonstrated substantial harm, though the RECOVERY trial have shown some evidence of harm looking at death and ventilation as a composite outcome.^27^ While hydroxychloroquine has been approved for use in treatment of RA and SLE for many years, recent evidence has suggested potential short-term harms (when co-prescribed with azithromycin).^28^ While we were underpowered to investigate co-medication with azithromycin, results of our study showed no evidence of an association with mortality from COVID-19 or other causes. This suggests justification to continue trials of hydroxychloroquine for prevention of COVID-19 to confirm our findings from observational data.

### Strengths and limitations

The greatest strengths of this study were the statistical power and the detailed longitudinal, routinely collected data to ascertain routine hydroxychloroquine use prior to the outbreak of COVID-19 in England. We were able to focus analyses on patients with indications for the use of hydroxychloroquine, a key component to mitigate confounding by indication in pharmacoepidemiological research of real-world data. Prior to starting analysis, we developed a DAG to identify a minimal set of covariates to adjust for the hypothesised confounding structure. We also fitted models adjusting for additional characteristics suggested as potentially important in consultation with clinicians. We performed informative sensitivity analyses including quantitative bias analyses to test key assumptions about missing data on bDMARD treatments. Lastly, the optimal timing^29^ and dose^30^ of hydroxychloroquine for therapeutic and prophylactic use for COVID-19 has been debated. Our population included regular users of hydroxychloroquine, in doses routinely used in clinical practice, with clarity that hydroxychloroquine administration occurred before exposure to SARS-CoV-2. Finally, we pre-specified our study protocol and have shared all analytical code.

We also recognise possible limitations. One is the risk of residual confounding by use of medications not prescribed in general practice, namely bDMARDs. While the majority of prescriptions in England are supplied by general practitioners in primary care, some medicines, including biologics, may be supplied by hospitals for various reasons including cost. We have advocated for these data to be more widely shared but, at present, they are not available.^31,32^ Although recent nationwide information on the prevalence of concomitant use of bDMARDs and hydroxychloroquine were unavailable, we demonstrated that our results were robust to a wide range of plausible assumptions about the use of these drugs and their potential relationship with COVID-19 mortality in quantitative bias analysis. A further potential source of confounding is severity of rheumatological disease, which is not captured in primary care records. However, the addition of a number of chronic comorbidities and proxies for health status in extended adjustment did not alter our findings. Another important consideration is the potential for exposure misclassification, whereby people prescribed hydroxychloroquine were not taking it as directed. In some reportedly rare cases in March, local shortages of hydroxychloroquine may have occurred due to inappropriate stockpiling; however, the United Kingdom has not suffered major shortages during the COVID-19 outbreak.^33^ An additional limitation in primary care prescribing data is drug exposure misclassification, whereby people may not adhere to medications as directed. Finally, COVID-19 mortality as an outcome reflects the risk of exposure to and acquiring SARS-CoV-2 infection, as well as the risk of developing severe disease and subsequent death. We were not able to explore the risk of SARS-CoV-2 infection in the current study due to the lack of complete or representative testing data. However, if hydroxychloroquine had a strong protective effect on the risk of SARSCoV-2 infection, we would have expected to see this reflected in lower risk of COVID-19 mortality.

### Summary

We carried out a large study of patients who were prescribed hydroxychloroquine for its licensed purpose and followed them up to look for clear signals of benefit in mortality from COVID-19 and other causes. We found no evidence of benefit after adjusting for important differences in those who had received hydroxychloroquine. Completion of randomised trials for prevention of severe outcomes is warranted to confirm these observational findings. The use of hydroxychloroquine for prevention of COVID-19 mortality outside trial settings is currently not justified.

## Data Availability

All of the code used for data management and analyses is openly shared online for review and re-use (https://github.com/opensafely/hydroxychloroquine-research). All iterations of the pre-specified study protocol are archived with version control (https://github.com/opensafely/hydroxychloroquine-research/tree/master/protocol).

## Acknowledgements

We are very grateful for all the support received from the TPP Technical Operations team throughout this work; for generous assistance from the information governance and database teams at NHS England / NHSX.

## Ethics

This study was approved by the Health Research Authority (REC 20/LO/0651) and by the LSHTM Ethics Board (#21863).

## Guarantor

CR/LS/BG are guarantors.

## Contributorship

Contributions are as follows:

Conceptualization CR LS ID WD SE LT BG

Data curation BM CM CB SB DE PI JC JP FH SH

Formal Analysis CR KB JB

Funding acquisition LS BG

Information governance CB AM JP BG

Methodology CR ND WH BM CM KB JB AS WH LS ID WD SE LT BG

Disease category conceptualisation and codelists CR BM CM KB AS AJW CB SB AM HC DE KW PI RM HD HM JC HF JP SH ID WD SE LT

Ethics approval EW HC LS BG

Project administration CR BM CM CB SB AM HC LS ID SW LT BG

Resources LS BG

Software BM CM EH AJW CB SB DE PI JC FH SH

Supervision LS ID WD SE LT BG

Visualisation CR KB

Writing (original draft) CR ND ID SE LT

Writing (review & editing) CT ND BM CM KB JB AS WH RC AJW EW CB SB AM HC DE KW PI RM HD AYSW HM JC HF JP FH SH LS ID WD SE LT BG

## Reference

1. Joint Formulary Committee. British National Formulary - Hydroxychloroquine. Accessed July 13, 2018. https://bnf.nice.org.uk/drug/hydroxychloroquine-sulfate.html

2. Liu J, Cao R, Xu M, et al Hydroxychloroquine, a less toxic derivative of chloroquine, is effective in inhibiting SARS-CoV-2 infection in vitro. Cell Discov. 2020; 6: 16.

3. Yao X, Ye F, Zhang M, et al In Vitro Antiviral Activity and Projection of Optimized Dosing Design of Hydroxychloroquine for the Treatment of Severe Acute Respiratory Syndrome Coronavirus 2 (SARS-CoV-2). Clin Infect Dis. Published online March 9, 2020. doi: 10.1093/cid/ciaa237

4. Chen Z, Hu J, Zhang Z, et al Efficacy of hydroxychloroquine in patients with COVID-19: results of a randomized clinical trial. MedRxiv. Published online 2020. https://www.medrxiv.org/content/10.1101/2020.03.22.20040758v3.abstract

5. COVID-19 Clinical Trials Report Card: Chloroquine and Hydroxychloroquine - CEBM. CEBM. Accessed July 13, 2020. https://www.cebm.net/covid-19/covid-19-clinical-trials-report-cardchloroquine-and-hydroxychloroquine/

6. Mitjà O, Corbacho-Monné M, Ubals M, et al Hydroxychloroquine for Early Treatment of Adults with Mild Covid-19: A Randomized-Controlled Trial. Clin Infect Dis. Published online July 16, 2020. doi: 10.1093/cid/ciaa1009

7. World Health Organisation. “Solidarity” clinical trial for COVID-19 treatments. Accessed August 3, 2020. https://www.who.int/emergencies/diseases/novel-coronavirus-2019/global-researchon-novel-coronavirus-2019-ncov/solidarity-clinical-trial-for-covid-19-treatments

8. Skipper CP, Pastick KA, Engen NW, et al Hydroxychloroquine in Nonhospitalized Adults With Early COVID-19: A Randomized Trial. Ann Intern Med. Published online July 16, 2020. doi: 10.7326/M20-4207

9. Horby P, Lim WS, Emberson J, et al Effect of Dexamethasone in Hospitalized Patients with COVID-19: Preliminary Report. medRxiv. Published online June 22, 2020. doi: 10.1101/2020.06.22.20137273

10. Rosenberg ES, Dufort EM, Udo T, et al Association of Treatment With Hydroxychloroquine or Azithromycin With In-Hospital Mortality in Patients With COVID-19 in New York State. JAMA. Published online May 11, 2020. doi: 10.1001/jama.2020.8630

11. Magagnoli J, Narendran S, Pereira F, et al Outcomes of Hydroxychloroquine Usage in United States Veterans Hospitalized with COVID-19. Med. Published online June 5, 2020. doi: 10.1016/j.medj.2020.06.001

12. Mahévas M, Tran V-T, Roumier M, et al Clinical efficacy of hydroxychloroquine in patients with covid-19 pneumonia who require oxygen: observational comparative study using routine care data. BMJ. 2020; 369: m1844.

13. Hernandez AV, Roman YM, Pasupuleti V, Barboza JJ, White CM. Hydroxychloroquine or Chloroquine for Treatment or Prophylaxis of COVID-19: A Living Systematic Review. Ann Intern Med. Published online May 27, 2020. doi: 10.7326/M20-2496

14. Boulware DR, Pullen MF, Bangdiwala AS, et al A Randomized Trial of Hydroxychloroquine as Postexposure Prophylaxis for Covid-19. N Engl J Med. Published online June 3, 2020. doi: 10.1056/NEJMoa201663815

15. Williamson EJ, Walker AJ, Bhaskaran K, et al OpenSAFELY: factors associated with COVID-19 death in 17 million patients. Nature. Published online 2020: 1–11.

16. Coronavirus (COVID-19) Research Platform. NHS England. Accessed August 8, 2020. https://www.england.nhs.uk/contact-us/privacy-notice/how-we-use-your-information/covid-19-response/coronavirus-covid-19-research-platform/

17. Thomas SL, Edwards CJ, Smeeth L, Cooper C, Hall AJ. How accurate are diagnoses for rheumatoid arthritis and juvenile idiopathic arthritis in the general practice research database? Arthritis Rheum. 2008; 59(9): 1314-1321.

18. World Health Organisation. Emergency use ICD codes for COVID-19 disease outbreak. Published May 22, 2020. Accessed August 5, 2020. https://www.who.int/classifications/icd/covid19/en/

19. Pintilie M. Analysing and interpreting competing risk data. Stat Med. 2007; 26(6): 1360–1367.

20. Bhaskaran K, Smeeth L. What is the difference between missing completely at random and missing at random? Int J Epidemiol. 2014; 43(4): 1336–1339.

21. VanderWeele TJ. Unmeasured confounding and hazard scales: sensitivity analysis for total, direct, and indirect effects. Eur J Epidemiol. 2013; 28(2): 113–117.

22. Choy E, Taylor P, McAuliffe S, Roberts K, Sargeant I. Variation in the use of biologics in the management of rheumatoid arthritis across the UK. Curr Med Res Opin. 2012; 28(10): 1733–1741.

23. Kim H. The Roles of Biosimilars in Patient Access to Biologics in UK. Accessed August 11, 2020. https://www.england.nhs.uk/expo/wp-content/uploads/sites/18/2018/09/12.30-Improvingpatient-care-with-biologics-in-the-management-of-RA-Celltrion-Healthcare-P2C.pdf

24. Arshad S, Kilgore P, Chaudhry ZS, et al Treatment with hydroxychloroquine, azithromycin, and combination in patients hospitalized with COVID-19. Int J Infect Dis. 2020; 97: 396–403.

25. Lee TC, MacKenzie LJ, McDonald EG, Tong SYC. An observational cohort study of hydroxychloroquine and azithromycin for COVID-19: (Can’t Get No) Satisfaction. Int J Infect Dis. 2020; 98: 216–217.

26. Griffith G, Morris TT, Tudball M, et al Collider bias undermines our understanding of COVID-19 disease risk and severity. medRxiv. Published online 2020.

27. Horby P, Mafham M, Linsell L, et al Effect of Hydroxychloroquine in Hospitalized Patients with COVID-19: Preliminary results from a multi-centre, randomized, controlled trial. medRxiv. Published online 2020.

28. Lane JCE, Weaver J, Kostka K, et al Safety of hydroxychloroquine, alone and in combination with azithromycin, in light of rapid wide-spread use for COVID-19: a multinational, network cohort and self-controlled case series study. medRxiv. Published online 2020. https://www.medrxiv.org/content/medrxiv/early/2020/05/31/2020.04.08.20054551.full.pdf

29. Lee SH, Son H, Peck KR. Can post-exposure prophylaxis for COVID-19 be considered as an outbreak response strategy in long-term care hospitals? Int J Antimicrob Agents. 2020; 55(6): 105988.

30. Watson JA, Tarning J, Hoglund RM, et al Concentration-dependent mortality of chloroquine in overdose. Elife. 2020; 9. doi: 10.7554/eLife.5863116

31. Goldacre B, MacKenna B. The NHS deserves better use of hospital medicines data. BMJ. 2020; 370. doi: 10.1136/bmj.m2607

32. Matthews A, Donaldson LJ, Evans SJ, Langan SM. Safety of medicines delivered by homecare companies. BMJ. 2018; 361: k2201.

33. Dr Elizabeth Price, President, British Society for Rheumatology. Letter: Hydroxychloroquine is not in short supply. Financial Times. Published April 10, 2020. Accessed August 15, 2020. https://www.ft.com/content/87becd4c-7a52-11ea-9840-1b8019d9a987

